# Post-discharge health status and symptoms in patients with severe COVID-19

**DOI:** 10.1101/2020.08.11.20172742

**Authors:** Himali Weerahandi, Katherine A. Hochman, Emma Simon, Caroline Blaum, Joshua Chodosh, Emily Duan, Kira Garry, Tamara Kahan, Savannah Karmen-Tuohy, Hannah C. Karpel, Felicia Mendoza, Alexander M. Prete, Lindsey Quintana, Jennifer Rutishauser, Leticia Santos Martinez, Kanan Shah, Sneha Sharma, Elias Simon, Ana Stirniman, Leora I. Horwitz

## Abstract

**Background:** Little is known about long-term recovery from severe COVID-19 disease. Here, we characterize overall health, physical health and mental health of patients one month after discharge for severe COVID-19.

**Methods:** This was a prospective single health system observational cohort study of patients ≥18 years hospitalized with laboratory-confirmed COVID-19 disease who required at least 6 liters of oxygen during admission, had intact baseline cognitive and functional status and were discharged alive. Participants were enrolled between 30 and 40 days after discharge. Outcomes were elicited through validated survey instruments: the PROMIS^®^ Dyspnea Characteristics and PROMIS^®^ Global Health-10.

**Results:** A total of 161 patients (40.6% of eligible) were enrolled; 152 (38.3%) completed the survey. Median age was 62 years (interquartile range [IQR], 50-67); 57 (37%) were female. Overall, 113/152 (74%) participants reported shortness of breath within the prior week (median score 3 out of 10 [IQR 0-5]), vs. 47/152 (31%) pre-COVID-19 infection (0, IQR 0-1), p<0.001. Participants also rated their physical health and mental health as worse in their post-COVID state (43.8, standard deviation 9.3; mental health 47.3, SD 9.3) compared to their pre-COVID state, (54.3, SD 9.3; 54.3, SD 7.8, respectively), both p <0.001. A total of 52/148 (35.1%) patients without pre-COVID oxygen requirements needed home oxygen after hospital discharge; 20/148 (13.5%) reported still using oxygen at time of survey.

**Conclusions:** Patients with severe COVID-19 disease typically experience sequelae affecting their respiratory status, physical health and mental health for at least several weeks after hospital discharge.

## Introduction

The body of literature on the inpatient course of illness of COVID-19 has rapidly grown over the past several months; however, little is known about the long-term recovery from severe COVID-19 disease.^1^ COVID-19 is characterized by an unusual degree of hypoxia among hospitalized patients, reports of delirium and encephalopathy, hypercoagulability and a much higher intubation and death rate than comparable viral pneumonias.^2-9^ The extent to which impacts on health persist after hospital discharge, however, is uncertain.

Anecdotal reports have described onset of pulmonary fibrosis in severely ill patients,^5^ which would be expected to cause long-term disability; however, only one study has comprehensively assessed duration and severity of dyspnea after recovery.^1^ This study of 143 patients in Italy found that 43% of patients reported persistent dyspnea an average of 60 days after symptom onset. Moreover, patients requiring mechanical ventilation have experienced longer periods of intubation^5^ than is typical for infectious pneumonia, increasing risk for muscle atrophy, delirium, pulmonary damage and long-term disability.^10^ Finally, anecdotes of neurologic manifestations of COVID-19 disease even without mechanical ventilation, such as encephalopathy and confusion, have been reported.^7,11^ Whether these manifestations persist post-recovery is also unknown.

Studying the long-term sequelae of COVID-19 disease is critical for understanding the full natural history of disease, accurately predicting the cumulative impact of disease beyond hospitalization and mortality, and determining whether inpatient or post discharge pulmonary rehabilitation should be considered. Here, we characterize overall health status and the physical and mental health of patients discharged home after severe COVID-19.

## Methods

### Study Cohort

This was a prospective single health system observational cohort study conducted at NYU Langone Health. Eligible patients were those 18 years and older who required at least 6 liters of oxygen at any point during a hospitalization for laboratory-confirmed COVID-19, who were discharged alive to either home or a facility after April 15, 2020, and were still alive at the time of study contact. We excluded patients with communication impairment or baseline dementia. This was determined by chart review or if upon consent for this study, the patient was unable to articulate the purpose of this study and what would be required of them to participate. We also excluded patients discharged to hospice, patients who resided in long-term care pre-hospitalization, patients fully dependent in activities of daily living pre-hospitalization, and patients that opted out of research.

We used the electronic health record to identify consecutive discharges within the prior 30-40 days. Trained study personnel manually reviewed each patient’s chart to verify eligibility. Eligible patients were then called to solicit interest in the study. Interested patients were consented and enrolled over the phone. After informed consent was obtained, the study survey (Appendix) was either conducted by study personnel with the patient over the phone or independently online by the patient, per the patient’s preference. If the patient was not English speaking, the survey was conducted over the phone with interpretation services. NYU Grossman School of Medicine’s Institutional Review Board approved the protocol.

### Instruments

Data were collected and managed using REDCap electronic data capture tools^12^ hosted at NYU Langone Health in order to minimize missing inputs and allow for real-time data validation and quality control. The survey was designed to reinforce, but not require, responses to all items. Missing individual item responses were not imputed other than as described below for the PROMIS^®^ Dyspnea Characteristics instrument.

Outcomes were elicited through validated PROMIS^®^ (Patient-Reported Outcomes Measurement Information System) survey instruments (Appendix). PROMIS^®^ is a program developed by the National Institutes of Health that provides rigorous measures of patient-reported outcomes for use in clinical studies in patients and in the general population.^13^ It affords comparability, reliability and validity, flexibility, and inclusiveness across a range of patients and diagnoses.^14^

To evaluate the degree of residual pulmonary impairment, we used the PROMIS^®^ Dyspnea Characteristics instrument (Appendix).^15^ This is a five-item instrument that assesses quantitative and qualitative descriptions of a person’s experience of dyspnea. The first four items use a 0-10 numeric rating scale (where 0 represents no shortness of breath and 10 represents the worst possible shortness of breath) and the last item uses a five-point Likert scale. The first item asks participants to rate their shortness of breath in general. If the participant has no shortness of breath, the instrument stops there and items 2-3 are assigned a score of 0, and item 5 is assigned a score of 1. If the participant reports shortness of breath, we asked the remaining four items, which address the intensity, frequency, duration and severity of dyspnea. This instrument was administered twice during the survey: once asking patients to reflect on their shortness of breath in the last seven days and then again to reflect on their shortness of breath before they had COVID-19. Items in this instrument were scored individually.

To evaluate overall health status and mental health, we used the PROMIS^®^ Global Health-10 instrument (Appendix).^16^ This is a 10-item standardized psychometric instrument that measures an individual’s overall physical, mental, and social health. Each item is converted to a five-point scale, in which higher scores indicate better health. Physical and mental health summary scores are produced using four items each. Raw scores for these domains are converted to normed t-scores. These scores are standardized such that a score of 50 (10) represents the mean (standard deviation) for the United States general population. The remaining two items are on general health and how well social activities are carried out; these items are scored individually. As with the PROMIS^®^ Dyspnea Characteristics instrument, we administered the Global Health-10 instrument twice, first asking patients to reflect on how they felt in the past seven days and then again to reflect on how they felt before they became ill with COVID-19.

### Statistical Analyses

We used descriptive statistics to illustrate patient characteristics. To make bivariate comparisons of symptoms before and after COVID-19 and account for paired data, we utilized the Wilcoxon signed rank test for ordinal data comparisons and the paired t-test for normally distributed linear data comparisons. Pairwise comparisons were made with patients who had responded to items reflecting on their experience both before and after COVID-19. For score-based analyses, patients with incomplete surveys were excluded from the calculation of total scores. We also conducted a subgroup analysis of patients who had shortness of breath prior to COVID-19, comparing their shortness of breath severity before and after COVID-19.

Data were analyzed using SAS version 9.4 (SAS Institute, Cary, NC). All analyses were two-tailed and we treated a p value of <0.05 as significant.

## Results

### Study Participation and Cohort Characteristics

We screened 538 COVID-19 discharges identified through the electronic health record. Of these, 137 were deemed ineligible (**Figure**). Of the remaining 397 eligible discharges, 135 could not be reached, 94 declined to participate, 7 were rehospitalized and too sick to participate, and 161 patients enrolled, of whom 152 completed the survey. Patients who were eligible but not enrolled were of similar age as those who consented (median age 64 years [interquartile range 57-72] vs 62 years [IQR 50-67]), had similar median length of stay (17 days [IQR 10-37] vs 18 days [IQR 10-31]), similar intensive care unit use (101 [43.4%] vs 73 [45.3%]), and similar need for mechanical ventilation (86 [36.9%] vs 59 [36.7]). However, patients who were eligible but not enrolled were less likely to speak English (153 [65.7%] vs 125 [77.6%]), more likely to need an interpreter (53 [23.0%] vs 30 [18.8%]), more likely to have Medicare insurance (103 [44.2%] vs 55 [34.2%]), less likely to have received extracorporeal membrane oxygenation during hospitalization (8 [3.4%] vs 13 [8.1%]), and more likely to be discharged home (133 [55.1%]) compared to consented patients (73 [45.3%]) (**Table 1**).

**Figure:**
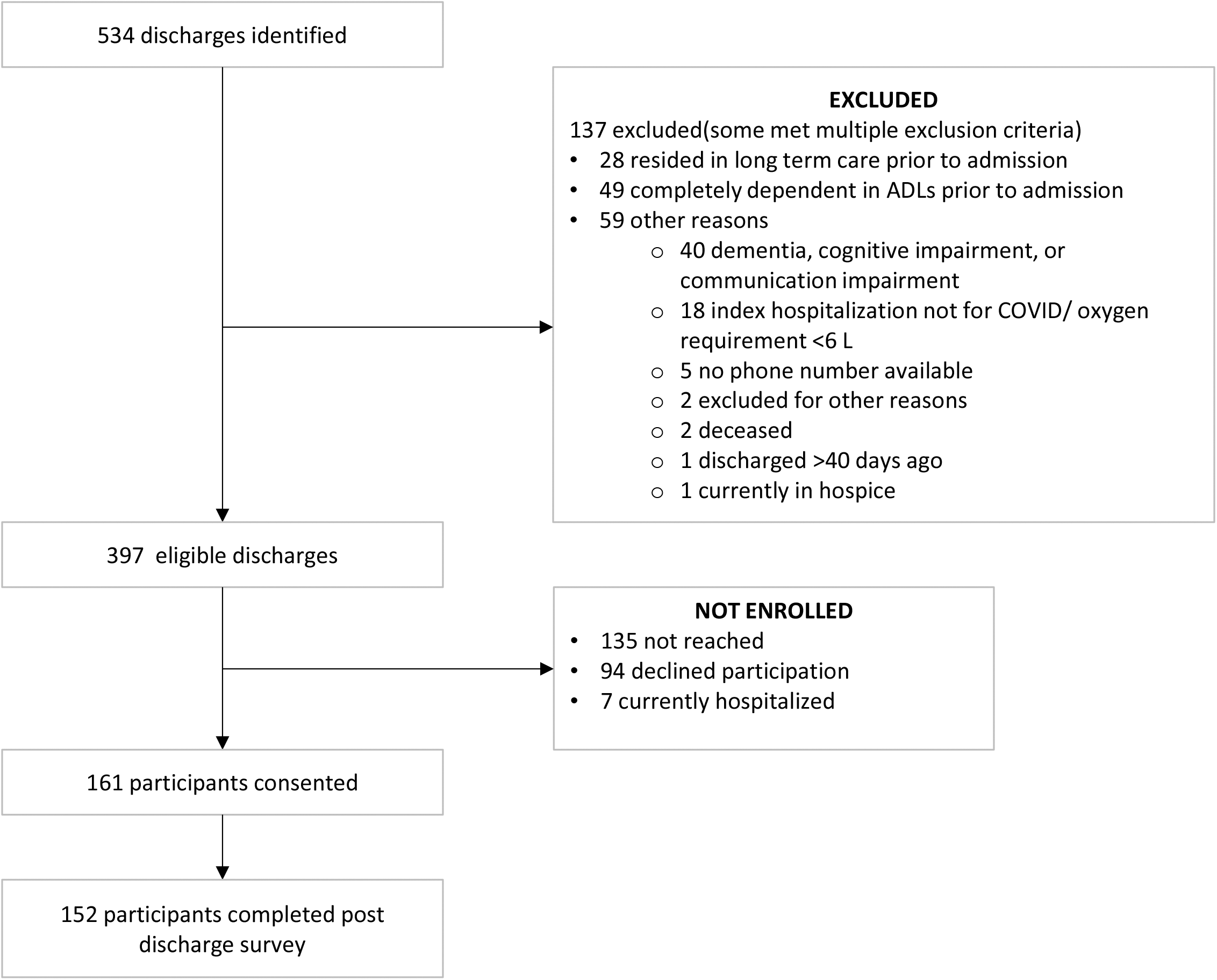
Flow diagram of enrolled participants

**Table 1:** Cohort Characteristics.

| **Characteristics** | **Consented**<br>**(N=161)**<br>**Median (IQR), or N (%)** | **Eligible but not enrolled**<br>**(N=233)**<br>**Median (IQR), or N (%)** |
| --- | --- | --- |
| Age (years), median (IQR) | 62 (50, 67) | 64 (57, 72) |
| Female | 60 (37.27) | 86 (36.91) |
| Race |  |  |
| White | 71 (44.1) | 87 (37.34) |
| Hispanic | 35 (21.74) | 74 (31.76) |
| Other/Multiracial | 14 (8.7) | 22 (9.44) |
| Asian | 16 (9.94) | 21 (9.01) |
| Black | 18 (11.18) | 22 (9.44) |
| Unknown | 7 (4.35) | 7 (3) |
| Language |  |  |
| English | 125 (77.64) | 153 (65.67) |
| Spanish | 26 (16.15) | 54 (23.18) |
| Russian | 4 (2.48) | 7 (3) |
| Bengali | 2 (1.24) | 1 (0.43) |
| Arabic | 1 (0.62) | 1 (0.43) |
| Cantonese | 1 (0.62) | 7 (3) |
| Mandarin | 1 (0.62) | 1 (0.43) |
| Haitian | 1 (0.62) | 1 (0.43) |
| Other | 0 (0) | 7 (3) |
| Unknown | 0 (0) | 1 (0.43) |
| Interpreter needed | 30 (18.75) | 53 (22.94) |
| Insurance |  |  |
| Commercial | 63 (39.13) | 66 (28.33) |
| Medicare | 55 (34.16) | 103 (44.21) |
| Medicaid | 39 (24.22) | 62 (26.61) |
| Unknown | 4 (2.48) | 2 (0.86) |
| Smoking status |  |  |
| Never | 94 (58.39) | 126 (54.08) |
| Former | 45 (27.95) | 57 (24.46) |
| Current | 4 (2.48) | 17 (7.3) |
| Unknown | 18 (11.18) | 33 (14.16) |
| Any chronic condition | 134 (83.23) | 189 (81.12) |
| Chronic Kidney Disease | 13 (8.07) | 38 (16.31) |
| Cancer | 12 (7.45) | 22 (9.44) |
| Coronary Artery Disease | 15 (9.32) | 41 (17.6) |
| Diabetes | 59 (36.65) | 100 (42.92) |
| Heart Failure | 8 (4.97) | 17 (7.3) |
| Hyperlipidemia | 75 (46.58) | 107 (45.92) |
| Hypertension | 97 (60.25) | 145 (62.23) |
| Asthma or chronic obstructive pulmonary disorder | 39 (24.22) | 32 (13.73) |
| BMI class |  |  |
| BMI <25 kg/m2 | 23 (14.29) | 67 (28.76) |
| BMI 25 to <30 kg/m2 | 49 (30.43) | 73 (31.33) |
| BMI 30 to <40 kg/m2 | 66 (40.99) | 71 (30.47) |
| BMI ≥ 40 kg/m2 | 22 (13.66) | 19 (8.15) |
| Unknown | 1 (0.62) | 3 (1.29) |
| Length of stay (days) | 18 (10, 31) | 17 (10, 37) |
| Intensive care unit stay | 73 (45.34) | 101 (43.35) |
| Type of respiratory support |  |  |
| Mechanical ventilation | 59 (36.65) | 86 (36.91) |
| High flow | 33 (20.5) | 39 (16.74) |
| Received ECMO | 13 (8.07) | 8 (3.43) |
| Received tocilizumab | 62 (38.51) | 74 (31.76) |
| Received corticosteroids | 21 (13.04) | 33 (14.16) |
| First D-dimer, median (IQR) | 404 (267, 650) | 505 (294.5, 959) |
| First C-reactive protein, median (IQR) | 140.7 (83.97, 190.1) | 145 (87.2, 214) |
| First RT-PCR cycle threshold, median (IQR) | 32.2 (26.9, 36.9) | 32.55 (27.7, 37.8) |
| Discharged to? |  |  |
| Home | 73 (45.34) | 133 (57.08) |
| Skilled nursing facility /rehab | 47 (29.19) | 73 (31.33) |
| Lower acuity inpatient setting | 1 (0.62) | 1 (0.43) |
| Unknown | 40 (24.84) | 26 (11.16) |
|  | **Completed Surveys**<br>**(N=152)**<br>**Median (IQR)** |  |
| Lives with |  |  |
| Spouse | 93 (57.8%) | N/A |
| Child(ren) | 79 (49.1%) | N/A |
| Grandchild(ren) | 10 (6.2%) | N/A |
| Other | 18 (11.2%) | N/A |
| Alone | 30 (19.7%) | N/A |
| Since hospital discharge... |  |  |
| Seen a medical professional | 137 (85.1%) | N/A |
| Received visiting nurse care | 88 (54.7%) | N/A |
| Had visiting nurse services prior to hospitalization | 1 (1.1%) | N/A |
| Received home health aide services | 27 (16.8%) | N/A |
| Had home health aide services prior to hospitalization | 5 (18.5%) | N/A |
| Had an ED visit | 9 (5.6%) | N/A |
| Were readmitted | 3 (1.9%) | N/A |

IQR: Interquartile range; BMI: Body mass index; ECMO: Extracorporeal membrane oxygenation

Participants completed surveys at a median of 37 days (range 30-43) after hospital discharge (equivalent to a median of 55 days (range 38-95) after hospital admission). Of those who completed surveys, 56/152 (36.8%) required home oxygen on hospital discharge, of whom 52 (92.9%) reported home oxygen as a new requirement; 20/148 (13.5%) participants previously not oxygen-dependent reported still requiring oxygen at the time of survey. For those with new current oxygen use, most (16/20, 80%) reported needing 1-2 liters per minute of oxygen.

Most participants (137, 90.1%) had seen a medical professional since discharge. Slightly more than half received visiting nurse services after hospital discharge; only one person reported having these services prior to hospitalization. In contrast, only 27 (17.8%) reported receiving home health aide services after hospital discharge; 5 (18.5%) participants in this group reported HHA services prior to hospitalization. Only 9 (5.9%) reported an ED visit after hospital discharge, and 3 (2.0%) reported hospital readmission.

### Dyspnea Outcomes

At the time of survey, a total of 113 (74.3%) participants reported some shortness of breath (median score 3 out of 10, IQR 0-5), compared to only 47 (30.9%) pre-COVID-19 infection (0, IQR 0-1), p<0.001. For those that did have shortness of breath prior to COVID-19, intensity, frequency, and duration of the shortness of breath worsened after COVID-19 (**Table 2**). In addition, more patients reported feeling short of breath “quite a bit” and “very much” in the past 7 days (18 [11.8%]) compared to before COVID-19 infection (4 [2.6%], p=0.028).

**Table 2:**
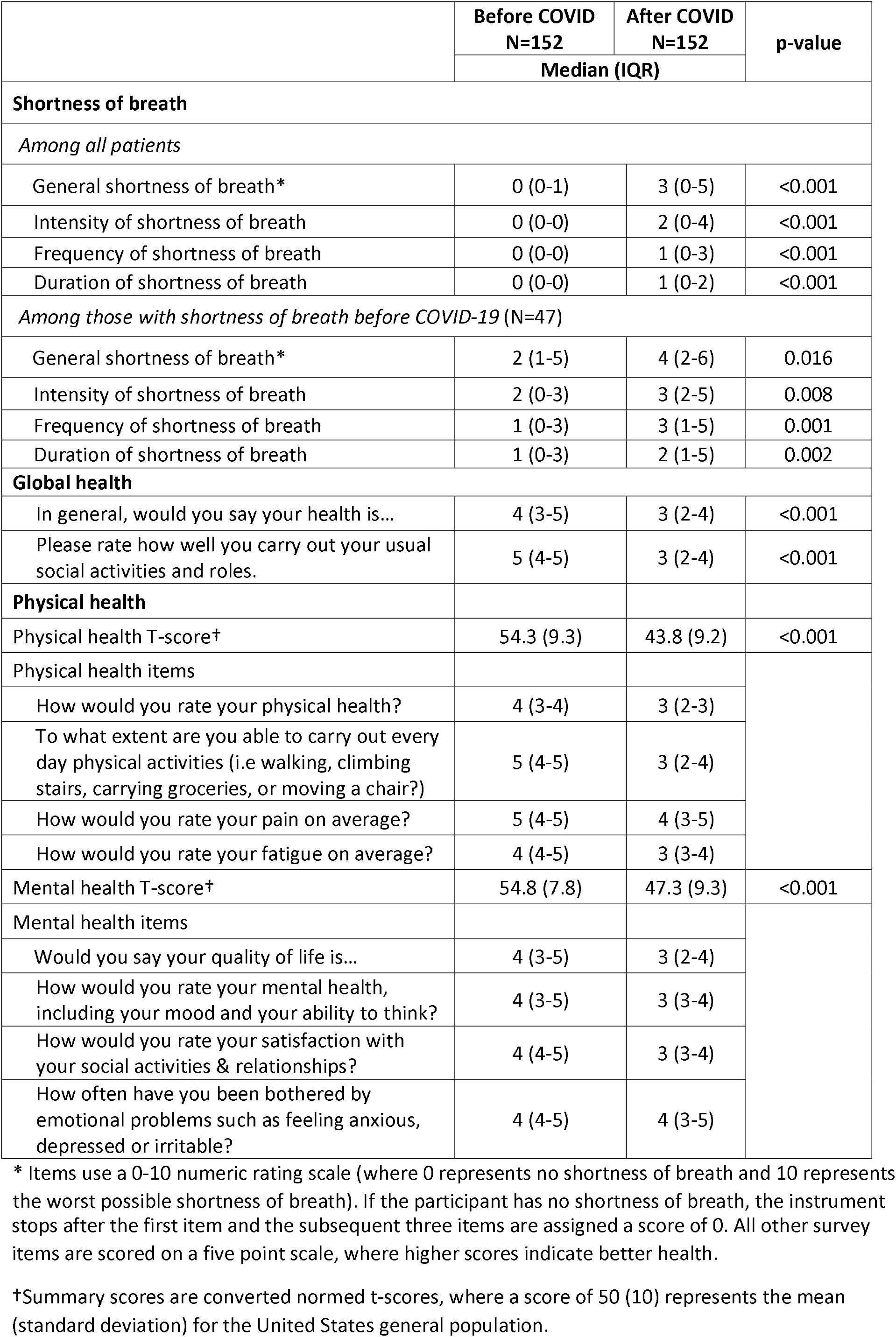
Outcomes.

### Overall Health, Physical Health and Mental Health

The PROMIS^®^ Global Health-10 instrument scores indicated worse general health after COVID-19 illness (3 out of 5, IQR 2-4) compared to baseline (4, IQR 3-5). Before COVID-19, participants’ summary t-scores in both the physical health and mental health domains were slightly above the United States mean of 50 (54.3, standard deviation 9.3; 54.3 SD 7.8, respectively). One month after COVID-19 infection, both scores were significantly lower (physical health: 43.8, SD 9.3; mental health 47.3, SD 9.3; p<0.001 for both). For the physical health score, this represents a decline of more than a full standard deviation. Patients also reported worsened ability to carry out social activities after COVID-19 (p<0.001). **Table 2** further details these outcomes.

## Discussion

In our prospective single health system observational cohort study examining post-discharge outcomes of patients hospitalized for severe COVID-19, we found that patients began with overall health slightly better than the United States average, and typically did not have shortness of breath prior to hospitalization for COVID-19. However, almost three quarters of patients reported persistent shortness of breath more than a month post-hospital discharge. For those who had shortness of breath prior to COVID-19, the intensity, frequency, and duration of the shortness of breath worsened after COVID-19. In addition, physical health and mental health were both rated as worse than baseline even one month after hospital discharge.

While anecdotal reports in the lay press have described patients with COVID-19 sequelae lasting for several weeks,^17-19^ our study confirms that this is the case for the great majority of patients who suffer from severe COVID-19 illness, not just isolated cases. The World Health Organization estimates that the median time from onset of disease to recovery of severe COVID19 is about 3-6 weeks, based on early data from China.^20^ However, three quarters of our patients had persistent dyspnea at 7-9 weeks after onset of disease (median of 55 days after hospital admission, which in turn typically occurs 5-10 days after symptom onset). Recent data from Italy examining post-discharge outcomes similarly indicate persistent dyspnea in 43% of patients an average of two months after symptom onset.^1^ Our cohort by design was more likely to have experienced intensive care or mechanical ventilation than the Italian cohort, likely explaining the higher prevalence of persistent dyspnea in our study.

Given the high incidence of intensive care unit stays and long hospitalizations associated with these cases, these patients are at risk for post-intensive care syndrome^21,22^ and post-hospital syndrome.^23^ This has serious implications for the ability to return to work, downstream effects on mental health due to sometimes drastic lifestyle and work capacity changes (most notably persistent shortness of breath), and the ability to engage in activities or hobbies enjoyed prior to COVID-19 illness.

Prior work examining post-discharge outcomes in acute respiratory distress syndrome (ARDS) demonstrate that while pulmonary function may recover, especially in relatively young patients, many experience reduced physical quality of life even at 5 years after their critical illness.^24-26^ Another study examining long-term outcomes in Influenza A (H7N9) survivors demonstrated improvement in pulmonary function and imaging findings during the first six months after hospital discharge but showed that impairment to quality of life persisted even at 2 years post-discharge.^27^

At this time, the permanence of reported symptoms for COVID-19 survivors is unclear; however, these early results are worrisome. Furthermore, we observed significant psychological distress in study participants, almost 20% of whom live alone. Given these findings, interventions after discharge may be needed to speed recovery.^28^ As shortness of breath may affect ability to complete activities of daily living, services to support these individuals during the post-discharge transition such as home health aide care should be arranged. Interventions for preventing and treating post-intensive care and post-hospital syndrome should also be considered. Pulmonary-specific interventions such as pulmonary rehabilitation should be explored.

### Strengths and Limitations

Our study provides a novel contribution characterizing early post-discharge outcomes in patients who have experienced severe COVID-19 disease. In addition, immigrant communities in New York City were hit particularly hard with COVID-19 and our study included patients from diverse backgrounds, with no language exclusions: nearly one in five patients in our study completed the survey through an interpreter. However, it has some limitations. Our survey does not include objective measures of pulmonary function. We compared current state to self-reported pre-COVID state, which may be subject to recall bias; however, this would not affect the post-COVID findings. We also specifically studied the experience of sicker patients---those who required hospitalization and at least 6 liters of oxygen during admission; our results are not generalizable to those with mild COVID-19. Of note, our results may underestimate the severity of post-discharge symptoms among those with severe COVID-19, as we excluded frail elders, people with dementia, and patients from or eligible for long-term care, who might be expected to have worse outcomes, and were unable to reach several patients who had been rehospitalized. Self-reported utilization outcomes such as readmission may also be underreported.^29^ Longer-term outcomes are still unknown. Generalizability may be limited by its single health system design; however, our health system does encompass multiple hospitals across urban and suburban settings, and our patient population was markedly diverse.

### Conclusion

The initial phase of the COVID-19 pandemic has taxed our healthcare system’s capacity to deliver acute and critical care. Prior work examining the long-term sequelae of severe ARDS has demonstrated deleterious effects on pulmonary function and health status even several years after disease onset.^25,26^ We found that survivors of severe COVID-19 experience shortness of breath, and worsened physical and mental health more than a month after hospital discharge. Whether these harms will persist is presently unknown. Regardless, it is clear that the individual and social impact of disease is more substantial than would be suggested by hospitalization and mortality rates alone. Given the volume of patients affected by this disease, plans to care for these patients longitudinally must be considered.

## Data Availability

Deidentified data may be available upon request to the authors.

## Acknowledgments

We would like to acknowledge Amy Bleasdale and Jacqueline L. Heath, MD for their contributions and Pacific Interpreters for providing free interpretation services for this study.

